# Association of baseline advanced HIV disease and dolutegravir versus non-dolutegravir regimen status with viral load suppression among patients on antiretroviral therapy in Tanzania

**DOI:** 10.64898/2026.03.19.26348804

**Authors:** Hamisi Amiri Dani, Prosper Njau, Raphael Z. Sangeda

**Affiliations:** Department of Pharmaceutical Microbiology, Muhimbili University of Health and Allied Sciences, Dar es Salaam, Tanzania; National AIDS, Sexually Transmitted Infections and Hepatitis Control Programme, Dodoma, Tanzania

**Keywords:** HIV, antiretroviral therapy, dolutegravir, advanced HIV disease, viral load suppression, Tanzania, routine data, cohort study

## Abstract

**Background:** Dolutegravir (DTG)-based regimens are currently the preferred first-line therapy in many HIV programs; however, the influence of baseline advanced HIV disease (AHD) on virologic outcomes in routine national data in the DTG era remains unclear.

**Methods:** We conducted a retrospective cohort analysis using routinely collected data from Tanzania’s National AIDS, STIs, and Hepatitis Control Programme (NASHCoP) database (2017-2021). A simple random sample of 50,000 patients was drawn from the de-duplicated national dataset, yielding 49,863 patients after data processing. The analytic cohort included 4,044 patients with baseline CD4 and endpoint viral load measurements. Viral load suppression was defined as <1000 copies/mL. Associations between baseline AHD, regimen status, and suppression were assessed using risk ratios and multivariable Poisson regression models, including an interaction term between AHD and DTG.

**Results:** Overall viral load suppression was 89.2% (3,607/4,044). Patients with baseline AHD had lower suppression than those without AHD (81.3% vs. 91.1%; RR 0.48, 95% CI 0.40-0.57). Suppression was higher among patients receiving DTG-based regimens than among those receiving non-DTG regimens (91.5% vs. 77.2%; RR 2.67, 95% CI 2.23-3.20). In the adjusted analysis, baseline AHD remained associated with reduced suppression (aRR 0.89, 95% CI 0.86-0.92), whereas DTG use was associated with improved suppression (aRR 1.15, 95% CI 1.10-1.20). A significant interaction between AHD and DTG was observed (aRR 1.40, 95% CI 1.20-1.63), indicating that the relative benefit of DTG was greater among patients with baseline AHD.

**Conclusions:** Although viral load suppression was high in this Tanzanian routine-care cohort, patients with baseline AHD had poorer outcomes. DTG-based regimens were associated with improved overall suppression, with a greater relative benefit among patients with advanced disease. These findings support the continued prioritization of DTG-based therapy and reinforce the importance of early diagnosis and targeted management of patients with AHD.

## Introduction

Despite substantial global progress in HIV diagnosis, treatment, and viral suppression, HIV remains a major public health challenge, with a disproportionate burden in sub-Saharan Africa [1,2]. The scale-up of antiretroviral therapy (ART) has markedly reduced HIV-related morbidity and mortality; however, treatment outcomes remain uneven across patient subgroups, particularly in resource-limited settings, where delayed presentation to care persists [3,4].

Advanced HIV disease (AHD) remains a critical concern in routine HIV care. Among adults and adolescents, AHD is defined as a CD4+ T-cell count of < 200 cells/µL or World Health Organization (WHO) clinical stage 3 or 4 disease [5]. Patients presenting with AHD are at increased risk of opportunistic infections, mortality, and suboptimal treatment outcomes. Despite the adoption of universal test-and-treat strategies, AHD remains common in sub-Saharan Africa [6–8]. Evidence from the region indicates that a substantial proportion of patients initiate care with severe immunosuppression, and these patients are less likely to achieve optimal virologic outcomes [9,10].

Tanzania has made considerable progress in expanding antiretroviral (ART) coverage and improving viral suppression, with national estimates approaching the UNAIDS 95-95-95 targets [2,11]. However, aggregate national indicators may mask important heterogeneity in treatment outcomes across patient groups, including those presenting with advanced disease.

The introduction of dolutegravir (DTG)-based regimens has transformed HIV treatment. The World Health Organization recommends DTG-based first-line ART owing to its high potency, favorable tolerability, and strong resistance barrier compared with older regimens [12]. Evidence from landmark randomized controlled trials—such as the SINGLE and FLAMINGO studies—alongside programmatic settings, has consistently demonstrated high rates of viral suppression with DTG-based therapies, often exceeding 90% [13–20]. More recent analyses using national routine care data from Tanzania have further demonstrated high viral suppression and highlighted the role of viral load monitoring and adherence patterns in shaping treatment outcomes [21–23]. These attributes have made DTG central to HIV treatment policy and program implementation in Tanzania and globally.

However, an important question remains regarding whether the benefits of DTG-based regimens are consistent across clinical subgroups, particularly among patients presenting with AHD. While severe immunosuppression at treatment initiation is known to be associated with poorer outcomes, evidence from national routine-care data examining the joint influence of baseline AHD and regimen type on viral suppression remains limited in the DTG era. Understanding this relationship is essential for identifying patients at risk of suboptimal outcomes and for optimizing treatment strategies in routine care settings.

This study assessed the association between baseline AHD and regimen status (DTG versus non-DTG) with endpoint viral load suppression among patients receiving antiretroviral therapy in Tanzania.

## Methods

### Study design and data source

We conducted a retrospective cohort analysis using routinely collected HIV care and treatment data from the National AIDS, STIs, and Hepatitis Control Programme (NASHCoP) master database in Tanzania. The dataset covered the period from 2017 to 2021 and included patient-and visit-level records from care and treatment clinics nationwide. Prior to the analysis, visit-level data were integrated and processed to construct a patient-level analytical dataset that combined laboratory records, including viral load and CD4 measurements, regimen history, visit dates, appointment information, and adherence-related indicators derived from longitudinal visit data. This aggregation ensured that all analyses were performed at the patient level.

### Sampling

From the de-duplicated national NASHCoP database, we selected a simple random sample of 50,000 unique patients without replacement using the sample() function in R, ensuring equal selection probability among eligible individuals. Following linkage and processing across multiple source datasets, the final working dataset contained 49,863 unique patients after excluding records with incomplete linkage or inconsistencies identified during aggregation.

### Study population and eligibility

The study population included individuals who received antiretroviral therapy (ART) in Tanzania during the study period. For the present analysis, we restricted the analytic cohort to patients with both a non-missing baseline CD4 measurement at ART initiation and a non-missing endpoint viral load result. Patients lacking either variable were excluded. The final analytic cohort, therefore, represents individuals with complete data on both baseline immunologic status and virologic outcomes.

### Variables

The primary outcome was viral load suppression, defined as a viral load <1000 copies/mL, in accordance with the World Health Organization and national programmatic definitions. The main exposures were baseline AHD, defined as a CD4+ T-cell count <200 cells/µL at ART initiation, and regimen status, categorized as DTG-based versus non-DTG-based regimens based on the final recorded regimen. DTG-based regimens were identified using predefined regimen codes corresponding to DTG-containing combinations, which were mapped to a binary indicator.

Additional variables included age at ART initiation, gender, follow-up duration, average adherence, and therapy change episodes, which served as a proxy for treatment complexity over time. Adherence measures were derived from longitudinal visit data using appointment and refill patterns and were incorporated into the patient-level dataset used for analysis.

### Data handling

Data were reviewed for consistency, plausibility, and completeness prior to analysis, including harmonization of variable formats and recoding of categorical variables. Derived variables, including viral load suppression and AHD status, were constructed using predefined thresholds. All analyses were conducted on de-identified data, and direct patient identifiers were removed prior to preparing the analytical dataset.

### Statistical analysis

We summarized the analytic cohort using counts and percentages for categorical variables and means with standard deviations for continuous variables. Viral load suppression was estimated overall and within strata, defined by baseline AHD and regimen status. Unadjusted associations were expressed as risk ratios with 95% confidence intervals. To examine joint effects, patients were categorized into four groups based on baseline AHD and regimen status, and suppression proportions were estimated within each stratum.

For the adjusted analyses, generalized linear models with a Poisson distribution and log link were used to estimate adjusted rate ratios for viral suppression. A main-effects model including baseline AHD, regimen status, and covariates was first fitted, followed by a second model that included an interaction term between baseline AHD and DTG regimen status. Robust variance estimation was applied to account for potential overdispersion. All analyses were performed using R (R Foundation for Statistical Computing, Vienna, Austria), and statistical significance was assessed at a two-sided alpha level of 0.05.

### Ethical considerations

Ethical clearance was obtained from the Muhimbili University of Health and Allied Sciences Directorate of Research, Publication, and Innovation (reference number DA.282/298/02L/504, February 25, 2025). Permission to access the data was granted by the NASHCoP. All data were anonymized prior to analysis.

## Results

### Study population and cohort derivation

A total of 49,863 patients were included in the sampled patient-level dataset, corresponding to an equal number of indexed records after de-duplication. Of these, 36,721 (73.6 %) patients had documented endpoint viral load measurements. Baseline CD4+ T cell counts were available for 5,243 patients (10.5%), and 4,044 (8.1%) patients had both baseline CD4+ T cell counts and endpoint viral load measurements and were included in the final analytic cohort *(Figure 1)*.

**Figure 1:**
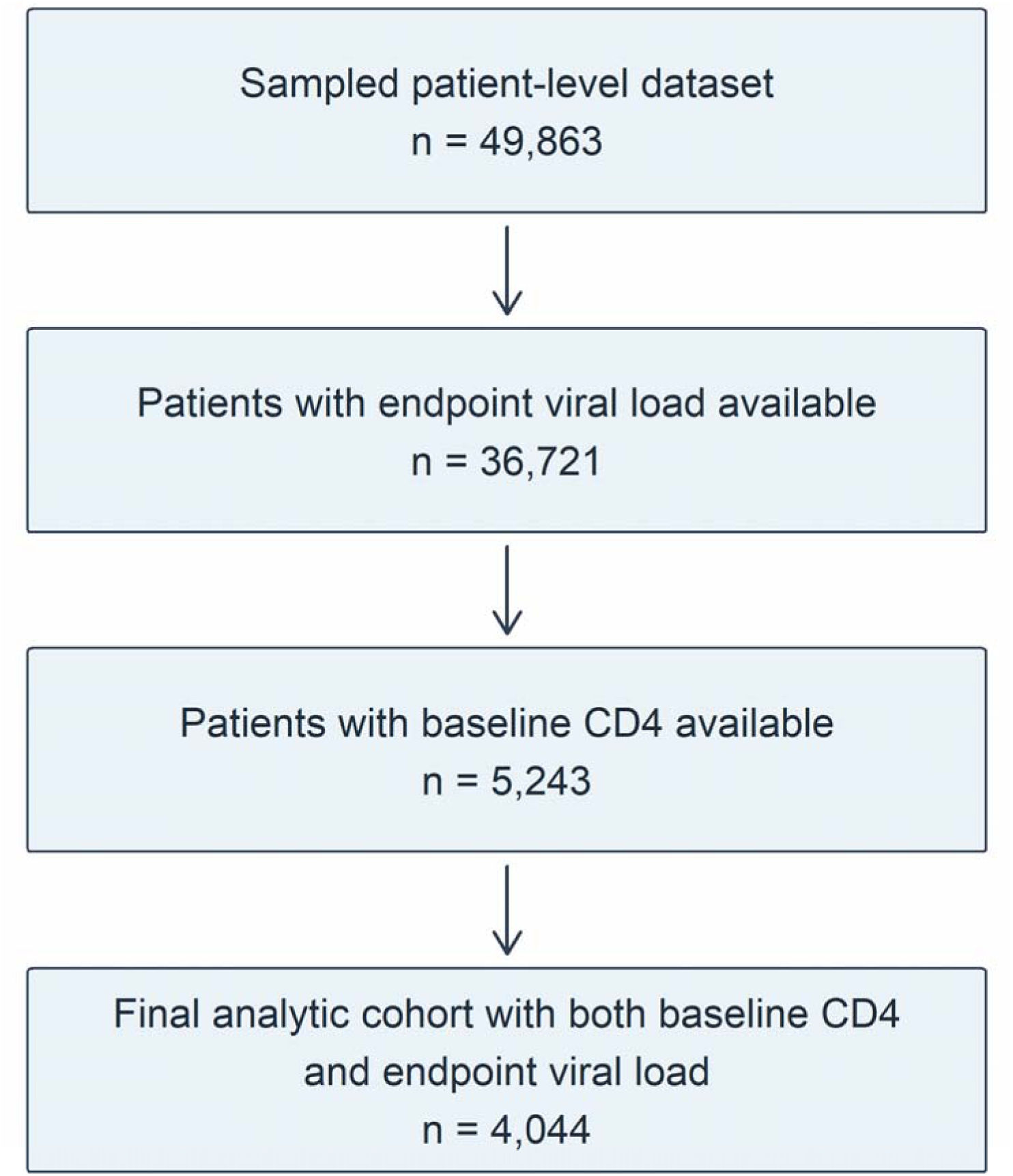
Cohort derivation flow diagram.

The analytic cohort was derived from the sampled patient-level dataset, which included endpoint viral load and baseline CD4 measurements, and the final number included in the analysis. In the analytical cohort, 3,607 patients achieved viral load suppression, corresponding to an overall suppression rate of 89.2%.

### Baseline characteristics of the analytic cohort

The mean age at ART initiation was 38.3 years (SD 13.3), increasing to 41.2 years (SD 13.5) at the end of follow-up. The mean follow-up duration was 1,262 days (SD 516). Patients had a mean of 2.7 therapy changes (SD 3.8) and 26 visits (SD 11.9) during the follow-up (Table 1).

**Table 1:** Baseline characteristics of the analytic cohort (N = 4,044)

| **Variable** | **Mean** | **SD** | **Min** | **Max** |
| --- | --- | --- | --- | --- |
| Age at ART start (years) | 38.3 | 13.3 | 0 | 94 |
| Age at end of follow-up (years) | 41.2 | 13.5 | 2 | 97 |
| Follow-up duration (days) | 1,262.2 | 516.4 | 0 | 1,824 |
| Therapy change count | 2.69 | 3.84 | 1 | 68 |
| Visit count | 25.97 | 11.92 | 1 | 104 |

Note: ART: Antiretroviral Therapy; SD: Standard Deviation.

### Viral load suppression by baseline advanced HIV disease status

Baseline AHD status influenced viral load suppression. Among patients without AHD (CD4 ≥200 cells/µL), 91.1% (2,963/3,252) achieved suppression, compared with 81.3% (644/792) among those with AHD.

This corresponded to a substantially lower likelihood of suppression among patients with AHD (RR 0.48, 95% CI 0.40-0.57) relative to those without AHD *(Table 2, Figure 2)*.

**Figure 2.**
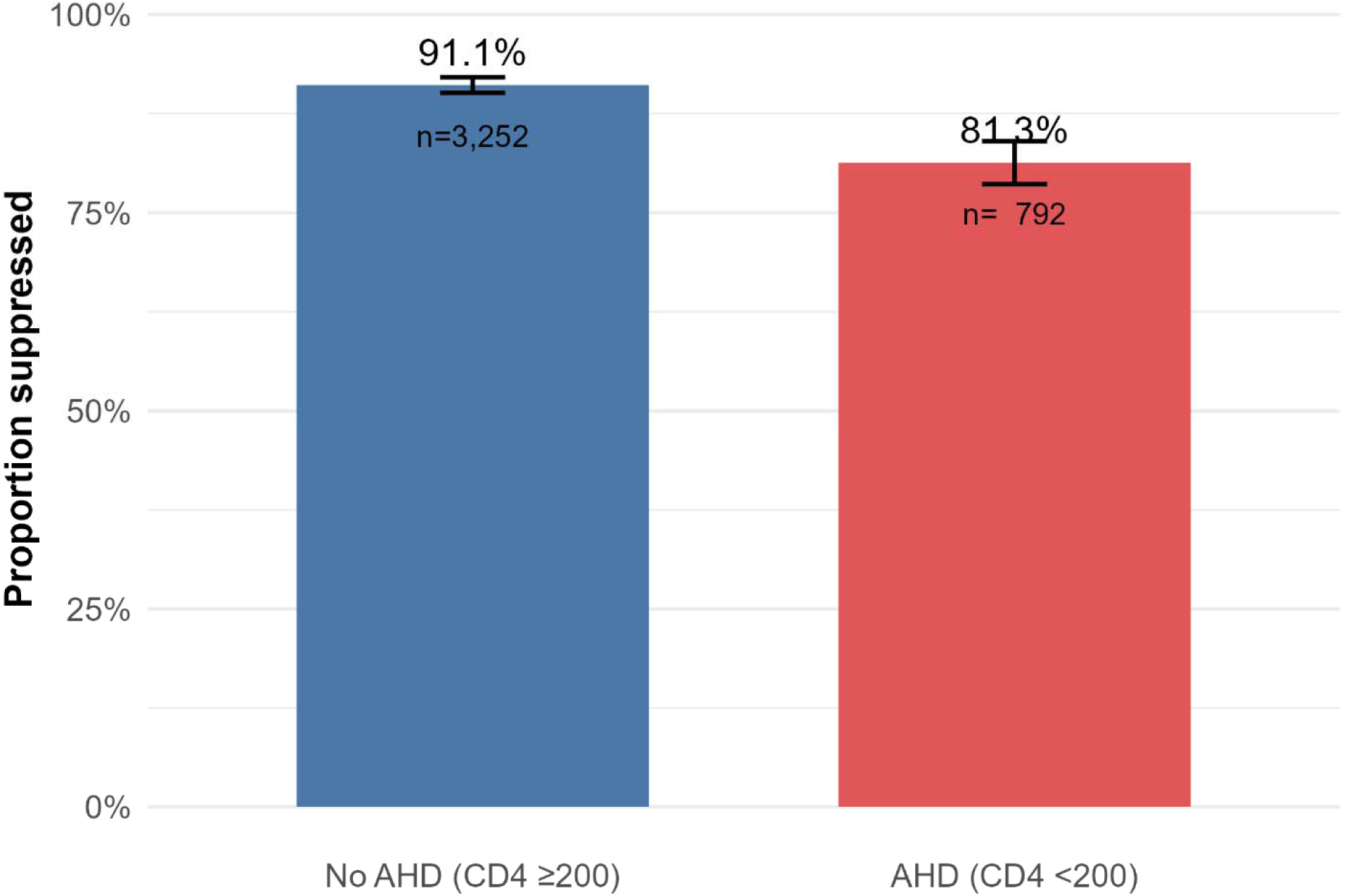
Endpoint viral load suppression according to baseline advanced HIV disease status.

**Table 2:**
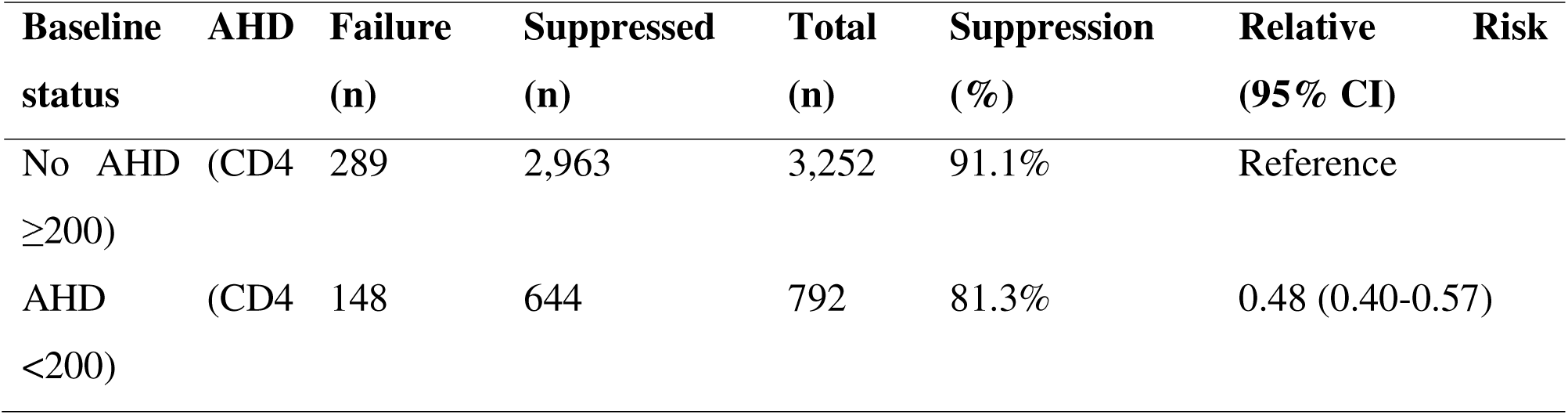
Viral load suppression by baseline advanced HIV disease (AHD)

Bar chart showing the proportion of patients with endpoint viral load suppression among those with and without baseline advanced HIV disease (AHD), defined as a CD4 count <200 cells/µL at ART initiation. Error bars represent 95% confidence intervals.

### Viral load suppression by final regimen status

Suppression rates were higher in patients receiving DTG-based regimens. Among patients on non-DTG regimens, 77.2% (498/645) achieved suppression, compared to 91.5% (3,109/3,399) among those on DTG-based regimens.

This corresponded to a markedly higher likelihood of suppression in patients treated with DTG (RR 2.67, 95% CI 2.23-3.20) than in those on non-DTG regimens *(Table 3, Figure 3)*.

**Figure 3:**
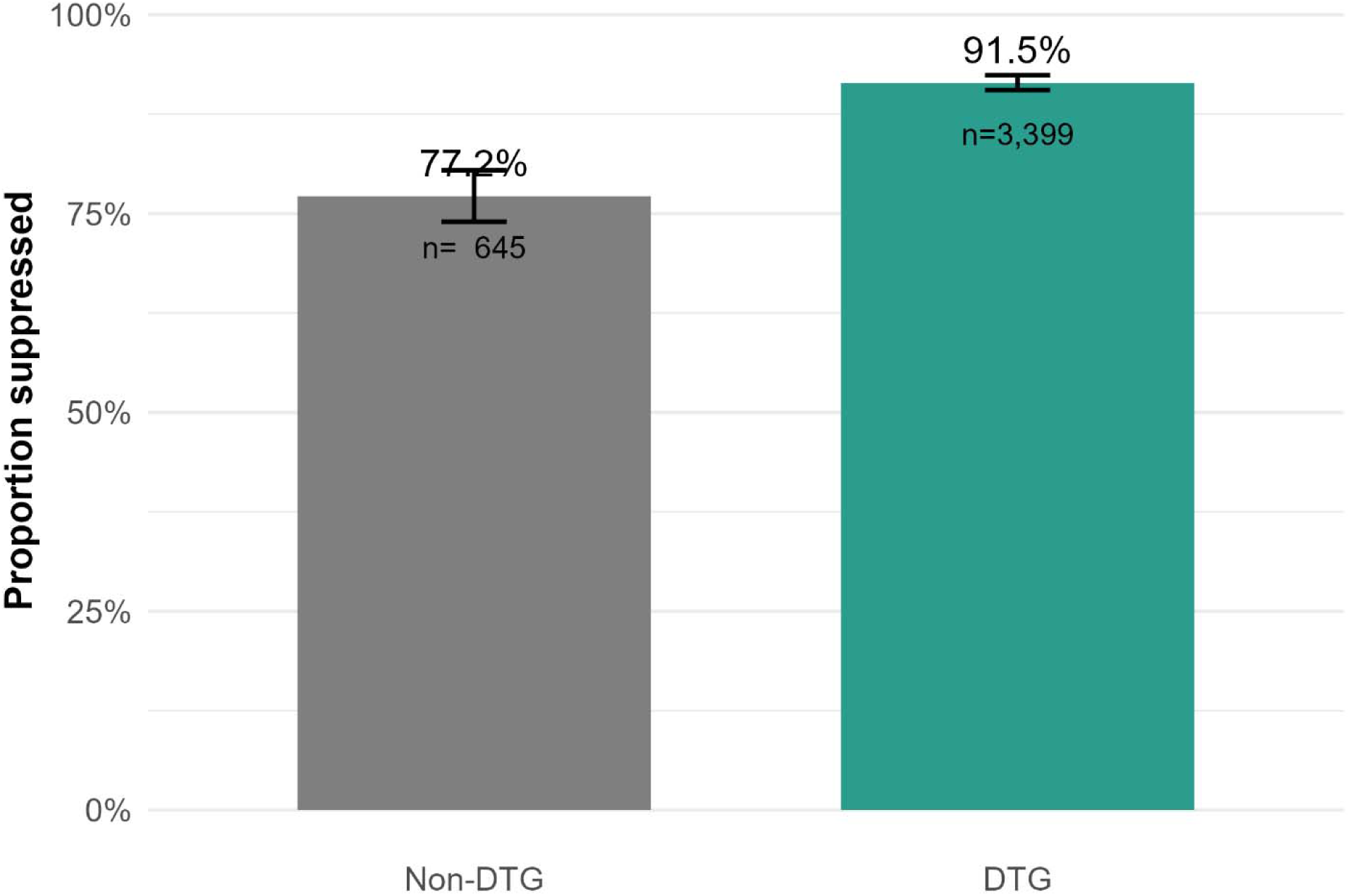
Endpoint viral load suppression according to final regimen status.

**Table 3:** Viral load suppression by final regimen status.

| **Regimen status** | **Failure (n)** | **Suppressed (n)** | **Total (n)** | **Suppression (%)** | **Relative Risk (95% CI)** |
| --- | --- | --- | --- | --- | --- |
| Non-DTG | 147 | 498 | 645 | 77.2% | Reference |
| DTG | 290 | 3,109 | 3,399 | 91.5% | 2.67 (2.23-3.20) |

The bar chart shows the proportion of patients with endpoint viral load suppression by final regimen status, categorized as DTG-based therapy versus non-DTG-based therapy. Error bars represent 95% confidence intervals.

### Viral load suppression across combined AHD and regimen strata

When stratified jointly according to baseline AHD and regimen status, clear gradients in viral suppression emerged (Table 4, Figure 4). Patients without AHD receiving DTG-based therapy exhibited the highest rates of suppression (92.5%), and those with AHD on the same regimen also maintained high suppression levels (87.2%).

**Figure 4.**
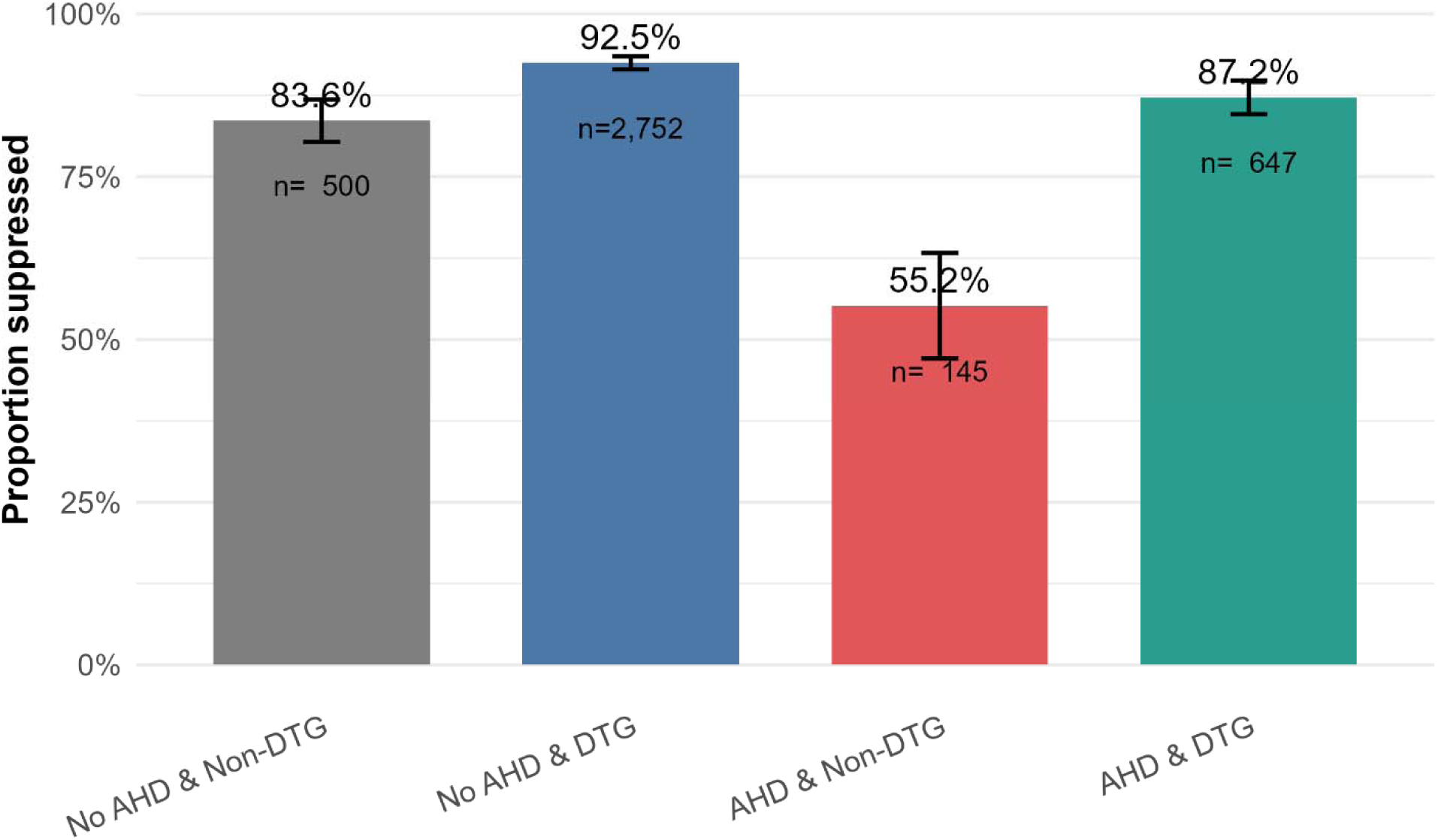
Endpoint viral load suppression across combined baseline advanced HIV disease and final regimen strata.

**Table 4:** Viral load suppression by combined baseline AHD and regimen status.

| Group | Total (n) | Suppressed (n) | Suppression (%) | 95% CI |
| --- | --- | --- | --- | --- |
| No AHD & Non-DTG | 500 | 418 | 83.6% | 80.4-86.8 |
| No AHD & DTG | 2,752 | 2,545 | 92.5% | 91.5-93.5 |
| AHD & Non-DTG | 145 | 80 | 55.2% | 47.1-63.3 |
| AHD & DTG | 647 | 564 | 87.2% | 84.6-89.7 |

In contrast, patients receiving non-DTG regimens experienced lower overall suppression. Within this non-DTG group, suppression was 83.6% for individuals without AHD, but fell substantially to 55.2% for those presenting with AHD.

Bar chart showing endpoint viral load suppression across the four groups defined by baseline advanced HIV disease (AHD) status and final regimen status: no AHD/non-DTG, no AHD/DTG, AHD/non-DTG, and AHD/DTG. Error bars represent 95% confidence intervals.

### Multivariable analysis of factors associated with viral load suppression-Main effects model

In the adjusted analysis, baseline AHD remained independently associated with a reduced likelihood of viral suppression (adjusted risk ratio [aRR] 0.89, 95% confidence interval [CI] 0.86-0.92, p < 0.001), whereas DTG-based regimens were associated with an increased likelihood of suppression (aRR 1.15, 95% CI 1.10-1.20, p < 0.001).

Higher adherence was also associated with improved suppression (aRR 1.004 per unit increase, p < 0.001), whereas increasing therapy change count was associated with a slight reduction in suppression (aRR 0.99, p = 0.007). Age at ART initiation showed a small positive association with suppression.

Gender and follow-up duration were not significant in the adjusted model *(Table 5)*.

**Table 5:** Adjusted model for factors associated with viral load suppression.

| **Variable** | **Adjusted Rate Ratio (aRR)** | **95% CI** | **p-value** |
| --- | --- | --- | --- |
| Baseline AHD (AHD vs No AHD) | 0.89 | 0.86-0.92 | <0.001 |
| DTG regimen (DTG vs Non-DTG) | 1.15 | 1.10-1.20 | <0.001 |
| Age at ART start (per year) | 1.002 | 1.002-1.003 | <0.001 |
| Female vs Male | 1.02 | 1.00-1.04 | 0.074 |
| Average adherence (per unit) | 1.004 | 1.002-1.005 | <0.001 |
| Follow-up duration (per day) | 1.000 | 1.000-1.000 | 0.054 |
| Therapy change count | 0.99 | 0.99-1.00 | 0.007 |

### Interaction model (AHD × DTG)

In the interaction model, a significant interaction between baseline AHD and DTG regimen status was observed (aRR 1.40, 95% CI 1.20-1.63, p < 0.001) (Table 6). Within this model, baseline AHD alone was strongly associated with reduced suppression (aRR 0.67), while DTG use remained associated with improved suppression (aRR 1.08).

**Table 6:** Adjusted model with interaction between baseline AHD and DTG regimen.

| Variable | Adjusted Rate Ratio (aRR) | 95% CI | p-value |
| --- | --- | --- | --- |
| Baseline AHD (AHD vs No AHD) | 0.67 | 0.58-0.78 | <0.001 |
| DTG regimen (DTG vs Non-DTG) | 1.08 | 1.04-1.13 | <0.001 |
| AHD × DTG interaction | 1.40 | 1.20-1.63 | <0.001 |
| Age at ART start (per year) | 1.002 | 1.002-1.003 | <0.001 |
| Female vs Male | 1.02 | 1.00-1.04 | 0.096 |
| Average adherence (per unit) | 1.004 | 1.002-1.005 | <0.001 |
| Follow-up duration (per day) | 1.000 | 1.000-1.000 | 0.088 |
| Therapy change count | 0.99 | 0.99-1.00 | 0.008 |

## Discussion

In this nationally representative routine-care cohort from Tanzania, overall endpoint viral load suppression was high (89.2%), indicating strong virologic performance among patients included in the analytic cohort. This estimate is broadly consistent with Tanzanian programmatic and survey-based evidence in the DTG era, which has reported high levels of viral suppression following the national scale-up of DTG-based regimens [11,15]. The findings suggest that among patients with available baseline CD4 and endpoint viral load measurements, most achieved favorable treatment outcomes during follow-up.

Despite this encouraging overall picture, patients presenting with baseline AHD experienced substantially poorer outcomes. Viral suppression was approximately 10 percentage points lower among patients with AHD than among those without AHD, and this difference persisted in adjusted analyses. This observation is clinically plausible and consistent with evidence from sub-Saharan Africa demonstrating that individuals initiating care with severe immunosuppression remain at elevated risk of virologic failure, opportunistic infections, and mortality, even in the context of expanded ART access [6,8,10]. These findings reinforce the continuing prognostic importance of baseline immunologic status in routine HIV care.

DTG-based regimens were associated with substantially greater viral suppression than non-DTG regimens. Viral suppression exceeded 90% among patients receiving DTG, whereas it remained below 80% among those receiving non-DTG regimens. This is consistent with the well-established advantages of DTG, including greater antiviral potency, rapid viral suppression, favorable tolerability, and a higher genetic barrier to resistance [12–14]. These findings also align with Tanzanian studies documenting improved outcomes following the national transition to DTG-based therapy [15,17,18]. In addition, these findings are consistent with recent analyses of national routine care data in Tanzania, which have demonstrated high viral suppression rates and have emphasized the importance of adherence and viral load monitoring in predicting treatment outcomes [21–23].

When baseline AHD and regimen status were considered jointly, a clear gradient in viral suppression emerged. Patients without AHD receiving DTG achieved the highest suppression, whereas those with AHD on non-DTG regimens had markedly poorer outcomes. Importantly, among patients with AHD, suppression was substantially higher in those receiving DTG compared with those on non-DTG regimens. In an adjusted analysis, a significant interaction between baseline AHD and DTG regimen status was observed, indicating that DTG had a more pronounced beneficial effect on viral suppression among patients with advanced disease. This suggests that DTG may partially mitigate the adverse impact of severe immunosuppression on treatment outcomes. Clinically, this is highly relevant, as patients with AHD represent a vulnerable subgroup that may benefit most from regimens with strong and durable virologic efficacy.

These findings have important programmatic implications. First, they support the continued prioritization of DTG-based regimens as the preferred backbone of first-line ART in routine care. Second, they highlighted the ongoing relevance of identifying patients with AHD at the time of treatment initiation. In the era of universal test-and-treat, baseline CD4 testing may receive less emphasis; however, our findings suggest that it remains valuable for clinical risk stratification and for identifying patients who may require closer monitoring and enhanced adherence support. Third, the results underscore the importance of earlier HIV diagnosis and linkage to care, which would reduce the proportion of patients presenting with advanced disease and improve overall treatment outcomes. Recent work using machine learning approaches on Tanzanian routine data has further demonstrated the predictive value of adherence measures for viral suppression, underscoring the importance of integrating adherence monitoring into routine HIV care [22].

This study has several strengths. It uses a nationally sampled routine-care dataset derived from the NASHCoP database, providing strong programmatic relevance and reflecting real-world clinical practice across Tanzania. The large sample size and inclusion of patients in diverse settings enhance the generalizability of the findings in the context of routine HIV care.

However, several limitations should be considered. First, the study relied on retrospectively collected routine data, which may be subject to missingness, recording inconsistencies, and measurement error. Second, the analytic cohort was restricted to patients with baseline CD4 and endpoint viral load data available, which may introduce selection bias and limit generalizability to all patients on ART. Third, several potentially important confounders, including comorbidities, socioeconomic factors, and detailed treatment history, were unavailable. Finally, although multivariable models were applied, residual confounding cannot be excluded.

Overall, this study demonstrates that viral load suppression in Tanzania is high in routine care, but that patients with baseline AHD remain at a disadvantage. DTG-based regimens are associated with improved suppression overall, and their benefits appear particularly important among patients with advanced disease. These findings support earlier HIV diagnosis and sustained DTG-based treatment scale-up, while highlighting the need for targeted management of patients presenting with AHD.

## Conclusion

In this nationally representative routine-care cohort in Tanzania, overall viral load suppression was high. However, suppression remained lower among patients with baseline AHD. DTG-based regimens were associated with substantially greater suppression than non-DTG regimens, and this benefit was more pronounced among patients with AHD. These findings support the continued prioritization of DTG-based therapy, alongside sustained efforts to promote early diagnosis, baseline CD4 assessment, and targeted management of patients presenting with AHD.

## Recommendations

Programs should continue to prioritize DTG-based regimens as the preferred ART backbone while strengthening efforts to identify patients presenting with AHD. Baseline CD4+ T-cell testing remains useful for clinical risk stratification, even in the test-and-treat era. Expanded HIV testing, earlier linkage to care, and more focused monitoring of patients with baseline AHD may further improve viral suppression outcomes.

## Declarations

### Author contributions

HAD contributed to study drafting, literature review, descriptive interpretation, and initial report preparation. RZS and PN conceived the analytical direction, supervised the work, guided methodology and statistical interpretation, and critically revised the manuscript.

### Ethical approval

Ethical approval was obtained from the MUHAS Directorate of Research, Publication, and Innovation (reference number DA.282/298/02L/504, dated February 2025).

## Data availability

The data used in this study were derived from routine programmatic records held by the NASHCoP and are not publicly available.

## Funding

No specific funding was declared for this study.

## Conflict of interest

The authors declare no conflicts of interest.

## Notes

### Competing Interest Statement

The authors have declared no competing interest.

### Funding Statement

This study did not receive any funding

### Author Declarations

Ethical approval was granted by the Research Ethics Committee (REC) of the Muhimbili University of Health and Allied Sciences (MUHAS) in Dar es Salaam, Tanzania, under the Directorate of Research, Publication, and Innovation (reference number DA.282/298/02L/504, February 25, 2025). The REC waived the requirement for individual informed consent because this study involved a retrospective analysis of strictly de-identified, routinely collected programmatic data. Permission to access the database was granted by the National AIDS, STIs, and Hepatitis Control Programme (NASHCoP). All data were anonymized prior to analysis.

## References

1. Chen Y, Li A-D, Yang Y, Lu J, Xu Y, Ji X, et al. Global, regional and national burden of HIV/AIDS among individuals aged 15-79 from 1990 to 2021. AIDS Res Ther. 2025;22: 51. doi:10.1186/s12981-025-00745-5

2. UNAIDS. Global HIV & AIDS statistics - Fact sheet. 2024 [cited 22 Jan 2026]. Available: https://www.unaids.org/en/resources/fact-sheet

3. Farahani M, Mulinder H, Farahani A, Marlink R. Prevalence and distribution of non-AIDS causes of death among HIV-infected individuals receiving antiretroviral therapy: a systematic review and meta-analysis. Int J STD AIDS. 2017;28: 636–650. doi:10.1177/0956462416632428

4. Lailulo Y, Kitenge M, Jaffer S, Aluko O, Nyasulu PS. Factors associated with antiretroviral treatment failure among people living with HIV on antiretroviral therapy in resource-poor settings: a systematic review and metaanalysis. Syst Rev. 2020;9: 292. doi:10.1186/s13643-020-01524-1

5. World Health Organization. Guidelines for managing advanced HIV disease and rapid initiation of antiretroviral therapy. 2017 [cited 19 Mar 2026]. Available: https://www.who.int/publications/i/item/9789241550062

6. Carmona S, Bor J, Nattey C, Maughan-Brown B, Maskew M, Fox MP, et al. Persistent High Burden of Advanced HIV Disease Among Patients Seeking Care in South Africa’s National HIV Program: Data From a Nationwide Laboratory Cohort. Clinical Infectious Diseases. 2018;66: S111–S117. doi:10.1093/cid/ciy045

7. Kitenge MK, Fatti G, Eshun-Wilson I, Aluko O, Nyasulu P. Prevalence and trends of advanced HIV disease among antiretroviral therapy-naïve and antiretroviral therapy-experienced patients in South Africa between 2010-2021: a systematic review and meta-analysis. BMC Infect Dis. 2023;23: 549. doi:10.1186/s12879-023-08521-4

8. Stöger L, Katende A, Mapesi H, Kalinjuma A V, van Essen L, Klimkait T, et al. Persistent High Burden and Mortality Associated With Advanced HIV Disease in Rural Tanzania Despite Uptake of World Health Organization “Test and Treat” Guidelines. Open Forum Infect Dis. 2022;9. doi:10.1093/ofid/ofac611

9. Lerango TL, Markos T, Yehualeshet D, Kefyalew E, Lerango SL. Advanced HIV disease and its predictors among newly diagnosed PLHIV in the Gedeo zone, southern Ethiopia. McCreesh N, editor. PLoS One. 2024;19: e0310373. doi:10.1371/journal.pone.0310373

10. Kachingwe E, Mutanda N, Ntjikelane V, Benade M, Manganye M, Malala L, et al. Characteristics and six-month viral load suppression of clients presenting with advanced HIV disease in South Africa. Moyo S, editor. PLOS Global Public Health. 2025;5: e0004927. doi:10.1371/journal.pgph.0004927

11. United Republic of Tanzania. Tanzania HIV Impact Survey 2022-2023. 2024 [cited 19 Mar 2026]. Available: https://www.nbs.go.tz/nbs/takwimu/THIS2022-2023/THIS2022-2023_Summary_Sheet.pdf

12. World Health Organization. Updated recommendations on first-line and second-line antiretroviral regimens and post-exposure prophylaxis and recommendations on early infant diagnosis of HIV. 2019 [cited 19 Mar 2026]. Available: https://www.who.int/publications/i/item/WHO-CDS-HIV-18.51

13. Fantauzzi A, Turriziani O, Mezzaroma I. Potential benefit of dolutegravir once daily: efficacy and safety. HIV AIDS (Auckl). 2013;5: 29–40. doi:10.2147/HIV.S27765

14. Cruciani M, Parisi SG. Dolutegravir based antiretroviral therapy compared to other combined antiretroviral regimens for the treatment of HIV-infected naive patients: A systematic review and meta-analysis. De Socio GV, editor. PLoS One. 2019;14: e0222229. doi:10.1371/journal.pone.0222229

15. Kamori D, Barabona G, Maokola W, Rugemalila J, Mahiti M, Mizinduko M, et al. HIV viral suppression in the era of dolutegravir use: Findings from a national survey in Tanzania. Onwuamah CK, editor. PLoS One. 2024;19: e0307003. doi:10.1371/journal.pone.0307003

16. Bwire GM, Aiko BG, Mosha IH, Kilapilo MS, Mangara A, Kazonda P, et al. High viral suppression and detection of dolutegravir-resistance associated mutations in treatment-experienced Tanzanian adults living with HIV-1 in Dar es Salaam. Sci Rep. 2023;13: 20493. doi:10.1038/s41598-023-47795-1

17. Majula RT, Mweya CN. Brief communication: Long-term treatment outcomes of transitioning to dolutegravir-based ART from efavirenz in HIV study participants in Mbeya, Tanzania. AIDS Res Ther. 2024;21: 98. doi:10.1186/s12981-024-00662-z

18. Mutagonda RF, Mlyuka HJ, Maganda BA, Kamuhabwa AAR. Adherence, Effectiveness and Safety of Dolutegravir Based Antiretroviral Regimens among HIV Infected Children and Adolescents in Tanzania. Journal of the International Association of Providers of AIDS Care (JIAPAC). 2022;21. doi:10.1177/23259582221109613

19. Walmsley SL, Antela A, Clumeck N, Duiculescu D, Eberhard A, Gutiérrez F, et al. Dolutegravir plus Abacavir–Lamivudine for the Treatment of HIV-1 Infection. New England Journal of Medicine. 2013;369: 1807–1818. doi:10.1056/NEJMoa1215541

20. Clotet B, Feinberg J, van Lunzen J, Khuong-Josses M-A, Antinori A, Dumitru I, et al. Once-daily dolutegravir versus darunavir plus ritonavir in antiretroviral-naive adults with HIV-1 infection (FLAMINGO): 48 week results from the randomised open-label phase 3b study. The Lancet. 2014;383: 2222–2231. doi:10.1016/S0140-6736(14)60084-2

21. Mutagonda RF, Lugoba MD, Mwakyomo J, Sambu V, Musiba G, Mutayoba B, et al. Uptake and predictors of viral load testing and viral suppression among people receiving antiretroviral therapy in mainland Tanzania. 2026. pp. 1–27. doi:10.64898/2026.03.11.26348142

22. Lugoba MD, Sangeda RZ, Mutagonda RF, Mwakyomo J, Musiba G, Sambu V, et al. Leveraging Machine Learning Models and Pharmacy Refill Adherence as a Cost-Effective Proxy for Predicting HIV Viral Suppression during Antiretroviral Therapy in Resource-Limited Settings. 2026. doi:10.64898/2026.01.05.26343496

23. Mwavika ET, Kunambi PP, Masasi SJ, Lema N, Kamori D, Matee M. Prevalence, rate, and predictors of virologic failure among adult HIV-Infected clients on second-line antiretroviral therapy (ART) in Tanzania (2018–2020): a retrospective cohort study. Bull Natl Res Cent. 2024;48: 96. doi:10.1186/s42269-024-01248-5

